# A framework for studying multi-omic risk factors and their interplay: application to coronary artery disease

**DOI:** 10.1101/2025.04.01.25324976

**Authors:** Jiaqi Hu, Chen Lin, Hongyu Zhao, Andrew T. DeWan

## Abstract

**Background:** Transcriptome-wide and proteome-wide association studies (TWAS and PWAS) have identified risk genes across complex diseases. However, the contributions of proximal risk variants and cross-omic interplay remain to be understood.

**Methods:** We propose an integrative framework to characterize disease-associated transcripts and proteins, and apply it to coronary artery disease (CAD). We employed S-PrediXcan on a large-scale genome-wide association study (GWAS) for CAD, with prediction models from GTEx v8 whole blood tissue and the Atherosclerosis Risk in Communities (ARIC) plasma protein. Conditional analyses adjusting for nearby CAD risk variants were performed to retain significant associations. Genes identified by both TWAS and PWAS were subsequently examined for associations with CAD risk factors, and colocalization analyses were performed for expression quantitative trait loci (eQTLs) and protein QTLs (pQTLs).

**Results:** TWAS identified 294 genes, and PWAS identified 79 genes, with 10 genes common between two analyses: *CHMP2B, CLIC4, IL6R, MIF, MXRA7, NME2, NUDT5, PCSK9, TAGLN2*, and *WARS*. The predicted transcripts and proteins of these genes exhibited consistent associations with CAD risk factors, and colocalization tests revealed shared signals between eQTLs and pQTLs.

**Conclusion:** Our summary-level data framework enables the construction of multi-omic risk profiles for CAD, advancing the understanding of its genetic etiology through integrated transcriptomic and proteomic analyses.

## Introduction

Genetic factors play an important role in the development of chronic diseases. For example, approximately 40-50% of the risk of coronary artery disease (CAD) is attributable to genetic components^1^. Understanding genetic risk profiles is essential for advancing knowledge of disease etiology and improving strategies for prevention and treatment. Genome-wide association studies (GWASs) have identified over 100,000 single nucleotide polymorphism (SNP)-phenotype associations^2^. However, these findings fail to fully account for the estimated heritabilities of many diseases^3^, and most identified SNPs are located in non-coding regions^4^, presenting significant challenges for a comprehensive characterization of the genetic basis of diseases.

To enhance the power of identifying risk genes and to better elucidate the biological mechanisms underlying disease-associated SNPs, transcriptome- and proteome-wide association studies (TWASs and PWASs) have been proposed^5,6^. These studies employ prediction models to impute transcript or protein levels and assess their associations with diseases of interest^7,8^. For example, Li et al. conducted a TWAS using prediction models derived from nine tissue types and found 114 genes significantly associated with CAD, including 18 genes not colocalized with known GWAS loci. Their study further validated the causal roles of two novel genes, *RGS19* and *KPTN*, through mouse experiments^5^. More recently, Zhao et al. (2023) performed a similar analysis and reported an association between the predicted expression of *NBEAL1* and CAD. This gene exhibited differential expression across macrophages, plasma cells, and endothelial cells^6^, highlighting its potential role in CAD pathogenesis.

The PWAS framework has been applied to various diseases, including gout, where researchers identified IL1RN–a gene related to gout in PWAS–as a potential drug target^8^. Moreover, Mendelian randomization (MR) analyses have successfully identified plasma proteins, such as *FES* and *PCSK9*, as potentially causal for CAD^9^. These findings underscore the potential of extending current GWAS discoveries and providing mechanism-based insights into interpretations by integrating transcriptomic and proteomic data.

Despite these progresses, several research gaps remain unaddressed. First, the relationships between findings from TWAS or PWAS and their underlying GWAS signals have not been thoroughly investigated, raising the possibility that the identified associations may be influenced by nearby GWAS variants. Second, limited studies have simultaneously examined multiple omic layers, leaving critical gaps in understanding the interplay between different omic levels and the effects on disease development.

To address these research gaps, we developed a framework for analyzing the genetic risk profile of diseases from a multi-omic perspective and applied it to CAD. First, we performed TWAS and PWAS using summary statistics from large-scale GWAS to expand the list of genes associated with the disease. Second, to examine the relationship between TWAS/PWAS findings and GWAS signals, we conducted conditional analyses to ensure that nearby risk loci could not fully explain the identified associations. Finally, we compared the TWAS and PWAS results and conducted colocalization analyses to construct molecular pathways elucidating the genetic etiology of CAD.

## Methods

The GWAS summary statistics used in this study is available at https://www.cardiogramplusc4d.org/. The other data that support the findings of this study are available from the corresponding author upon reasonable request.

### GWAS summary statistics

We used the large-scale GWAS meta-analysis of predominantly European subjects conducted by Aragam et al. (2022)^10^. This meta-analysis included 181,522 CAD patients and 1,165,690 non-CAD controls, covering 20,073,070 genetic variants through an inverse-variance weighted approach. Detailed methods are described in the original study^10^. To ensure high-quality data, we performed additional quality control on the variants, including: (1) keeping only SNPs, (2) converting genomic coordinates to the GRCh38 reference build using LiftOver, and (3) matching the variants to those in the 1000 Genomes reference panel provided by S-PrediXcan. After these steps, 13,851,199 SNPs were retained for analysis.

### X-WAS

An overview of the methodology can be found in Figure 1. Associations between genetically predicted gene expression levels and disease risk were assessed using GWAS summary statistics and elastic net models from GTEx v8 whole blood tissue^11^.

**Figure 1.**
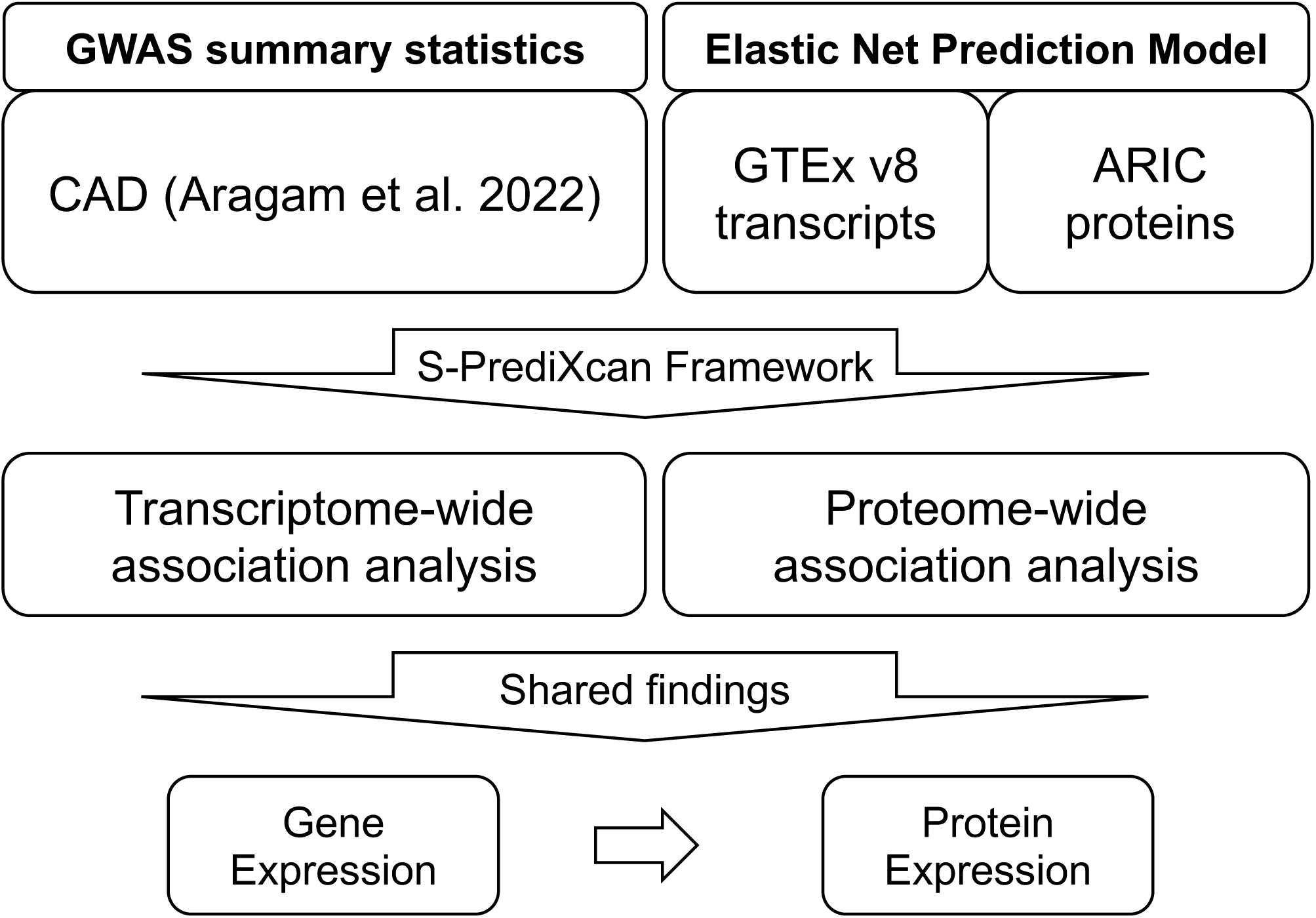
Flowchart of analysis. Overview of the methodology. Prediction models for gene and protein expression, along with large-scale GWAS, were used for TWAS and PWAS, respectively, via S-PrediXcan. Shared findings between TWAS and PWAS were identified.

These analyses were conducted using S-PrediXcan^7^, a summary-level method that infers relationships between genetically predicted gene expression and disease risk. Specifically, SNPs in the GWAS summary statistics were matched to genes based on established models, and Z-scores and p-values were calculated to test the null hypothesis that a given gene expression level was not associated with disease risk.

A similar framework was applied for protein expression (PWAS). Using GWAS summary statistics and elastic net prediction models for plasma protein expression levels from the Atherosclerosis Risk in Communities (ARIC) study European samples^8^, we inferred the associations between protein expression and disease risk through an extended version of S-PrediXcan designed for protein expression^8^.

To evaluate genomic inflation, Lambda 1,000 (λ_1000_) was calculated for both TWAS and PWAS results. This metric scales the genomic inflation factor to a population of 1,000 cases and 1,000 controls, using the formula: 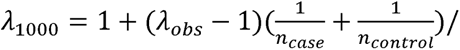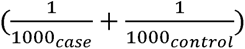, where λ_obs_ is the observed genomic inflation factor, and *n_case_* and *n_control_* are the numbers of cases and controls in the GWAS summary statistics, respectively. A λ_1000_ value close to 1.05 indicated a stable X-WAS analysis^12^.

### Conditional analysis

To determine whether the associations between gene or protein expression levels and disease risk were fully attributable to nearby GWAS risk SNPs, we conducted a conditional analysis on specific SNPs. First, index SNPs for CAD were identified using

LD clumping on GWAS summary statistics. Variants with p-values < 5×10^-6^ and R^2^ < 0.2 were retained. Next, we selected index variants located within ±1Mb of the genes of interest and compared these SNPs with GTEx V8 significant eQTLs for the corresponding genes, retaining only those that were not eQTLs. Conditional GWAS was then performed using GWAS and genotype data from 1000 Genomes European subjects^13^ through GWAS-COJO^14^. Following this step, X-WAS analyses were conducted. Genes with p-values greater than 0.05 were excluded from further analysis.

### Associations with risk factors

To investigate the functional relevance of the identified genes, we examined their associations with relevant risk factors in a cohort of unrelated White British subjects from the UK Biobank (UKB) (N=307,230). Transcript and protein levels were predicted using high-quality imputed genotype data and established models from GTEx and ARIC through PrediXcan^15^. The predicted omic levels were then standardized. We selected 15 phenotypes related to the three diseases of interest, including ever smoking, current smoking, hypertension (defined as an ICD-10 diagnosis, use of anti-hypertension medications, systolic blood pressure (SBP) ≥ 140 mm Hg, or diastolic blood pressure (DBP) ≥ 90 mm Hg), measured SBP, measured DBP, total cholesterol (TC), low-density lipoprotein (LDL), high-density lipoprotein (HDL), triglycerides (TG), body mass index (BMI), C-reactive protein (CRP), neutrophil-to-lymphocyte ratio (NLR), lymphocyte-to-monocyte ratio (LMR), platelet-to-lymphocyte ratio (PLR), and systemic immune-inflammation index (SII). Logistic regression analysis was performed for each disease, using the risk factors as exposure variables, while adjusting for age, sex, and the top 10 genetic principal components (PCs). To account for multiple comparisons, we applied study-specific false discovery rate (FDR) correction.

### Comparison with GWAS Catalog

TWAS and PWAS have greater power for identifying risk factors compared to traditional GWAS. To explore whether the genes identified in our analysis were previously reported in GWAS, we compared our findings with records in the GWAS Catalog^2^. We first extracted the SNP-CAD association results from the GWAS Catalog (https://www.ebi.ac.uk/gwas/). Variants located within ± 500 Kb of the gene of interest were selected. For genes without reported nearby SNPs, we performed an additional search in the GWAS Catalog for traits associated with these genes. Specifically, SNPs within the ± 500 Kb range of each gene were mapped, and the traits or diseases associated with these variants were queried using the R package ‘gwasrapidd’^16^.

### Sensitivity analysis

To maintain consistency with the PWAS tissue, we used gene expression prediction models developed from whole blood. To assess tissue specificity, and whether the identified associations were unique to whole blood, we selected coronary artery tissue for sensitivity analysis as it is closely related to CAD. We conducted TWAS with the prediction models from coronary artery tissue of GTEx v8 to evaluate how many of the findings remained nominally significant in this alternative tissue.

We conducted an additional sensitivity analysis to evaluate the robustness of our TWAS findings to cell-type specificity using the OneK1K single-cell RNA sequencing (scRNA-seq) data for eight major cell types (B cell, CD4 cell, CD8 cell, Dendritic cell, Monocyte cell, Natural killer cell, plasmablast cell, and other T cell)^17^.

An elastic net model was used to train the cell-type-specific TWAS prediction model for each cell type separately^18^. Genes were selected if their models met the quality thresholds (prediction R^2^ > 0.01 and p-value < 0.05). Finally, the S-PrediXcan framework was applied to the GWAS using the single-cell prediction models and genes with a p-value less than 0.05 were significant.

The robustness of our PWAS findings to the prediction models was assessed using protein data in the UKB. Directly measured protein levels from 29,422 unrelated White British subjects were analyzed for associations with prevalent CAD. CAD cases were defined using a combination of self-reported surveys and hospital records^19^. Logistic regression models were then developed, adjusting for age at recruitment, sex, and the top 10 genetic PCs, with a significance threshold set at 0.05.

### Integrate multi-omic risk factors

We compared the TWAS and PWAS results to identify genes for which both predicted transcript and protein levels were significantly associated with CAD. For these shared genes, the correlations between predicted transcript and protein levels were analyzed using UKB data.

Colocalization tests were conducted to investigate whether gene and protein expression shared the same genetic variants. SNPs identified as significant eQTLs in GTEx V8 whole blood tissue and pQTLs in the ARIC dataset (p-values below the Bonferroni correction threshold) were used. Colocalization tests were performed with the R package ‘coloc’^20^. Posterior probabilities (PPs) were calculated for five hypotheses, with a PP3 or PP4 value greater than 0.9 indicating a potential shared genetic mechanism between gene and protein expression.

### Statistical analysis

To address the increased risk of type I error due to multiple comparisons, we applied the FDR control method^21^ unless otherwise stated. Most of the analyses were conducted using R version 4.2.

## Results

### TWAS

We analyzed 7,249 predicted transcripts (Supplemental Figure 1A), identifying 402 significantly associated with CAD (p-value ≤ 2.76×10-3) (Supplemental Table 1). After conditional analysis, 108 predicted transcripts were excluded, leaving a final set of 294 predicted transcripts. The top five genes with the lowest p-values were *CARF, PSRC1, FES, LIPA,* and *GGCX*. Sensitivity analyses were performed to evaluate the robustness of our findings to tissue specificity and cell-type specificity. The coronary artery tissue prediction models only evaluated 125 genes among 294 genes, with 97 (77.6%) retaining nominal significance (Supplemental Table 1). Furthermore, 250 genes out of the 294 genes were mapped in the OneK1K data, and 1020 cell-type-specific prediction models were constructed after quality control, of which 192 exhibited nominally significant associations (p-value < 0.05) in at least one cell type (Supplemental Table 1). These analyses collectively support the robustness of our findings across different tissue and cell type contexts.

In addition, many of these transcripts were significantly associated with CAD risk factors, particularly lipids traits (Supplemental Table 2). For example, the predicted transcript of *LPL* was positively associated with HDL and negatively associated with triglyceride and LDL, consistent with its protective effects on CAD. Associations with hypertension and blood pressure measurements were also observed, such as the negative associations between *FES* and hypertension, SBP, and DBP.

Comparison with the GWAS catalog revealed that 95 of these genes had not been previously linked to CAD. However, SNPs near all 95 genes were associated with CAD risk factors (Supplemental Table 3), supporting their potential relevance to CAD. Some associations aligned with our findings; for example, SNPs near *AC017074.2* were associated with SBP in previous studies, and we identified significant associations between the predicted transcript of *AC017074.2* and both SBP and DBP. However, certain associations observed in the GWAS catalog were not identified in our risk factor analysis, suggesting that nearby SNPs may directly influence gene expressions.

### PWAS

Among 1,345 predicted proteins analyzed (Supplemental Figure 1B), 97 unique proteins were significantly associated with CAD risk (p-value ≤ 3.51×10-3; Supplemental Table 1). After conditional analysis, 21 proteins with p-values ≥ 0.05 were excluded, leaving 79 proteins for subsequent investigation (Supplemental Table 1). The top five proteins with the strongest significance were PCSK9, FN1, ANGPTL4, IL6R, and PDE5A. In a sensitivity analysis involving 30 directly measured proteins, 21 demonstrated nominal significance, thereby supporting the robustness of our prediction models (Supplemental Table 1).

Further analysis revealed that many predicted proteins were strongly associated with lipid traits, exhibiting extremely low p-values (Supplemental Table 2). For instance, *APOC1* and *APOC3*, members of the apolipoprotein C (APOC) gene family, were significantly associated with increased levels of LDL, total cholesterol, and triglycerides, as well as decreased levels of HDL. Both predicted proteins were positively associated with CAD, suggesting their effects on CAD might function via lipids metabolism pathways.

Additionally, we identified 22 genes not previously linked to CAD in the GWAS catalog. SNPs near these genes were associated with CAD risk factors (Supplemental Table 3). For example, SNPs near *ADK* were associated with leukocyte count, and its predicted protein expression was significantly associated with SII in the UKB. Conversely, some genes, such as *CTSF*, showed associations with CAD risk factors for nearby SNPs but no direct associations in our risk factor analysis. Similar to findings in the TWAS analysis, this suggests that nearby SNPs might not necessarily influence the protein expression directly.

### Interplay

We identified 10 genes shared between the TWAS and PWAS analyses (*CHMP2B, CLIC4, IL6R, MIF, MXRA7, NME2, NUDT5, PCSK9, TAGLN2*, and *WARS*) (Table 1).

**Table 1.**
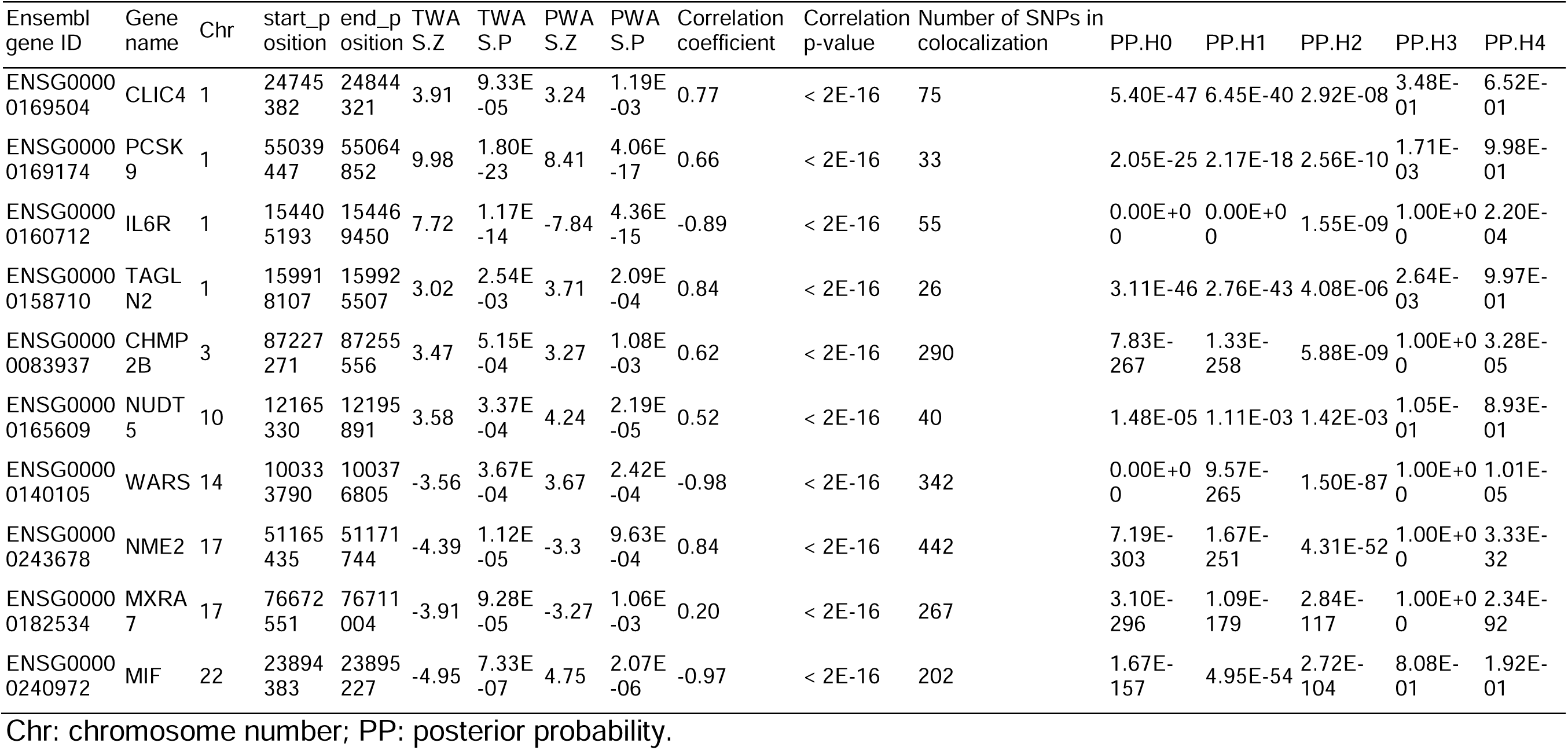
Ten shared genes between TWAS and PWAS.

| Ensembl<br>gene ID | Gene<br>name | Chr | start_p<br>osition | end_p<br>osition | TWA<br>S.Z | TWA<br>S.P | PWA<br>S.Z | PWA<br>S.P | Correlation<br>coefficient | Correlation<br>p-value | Number of SNPs in<br>colocalization | PP.H0 | PP.H1 | PP.H2 | PP.H3 | PP.H4 |
| --- | --- | --- | --- | --- | --- | --- | --- | --- | --- | --- | --- | --- | --- | --- | --- | --- |
| ENSG00000169504 | CLIC4 | 1 | 24745382 | 24844321 | 3.91 | 9.33E-05 | 3.24 | 1.19E-03 | 0.77 | < 2E-16 | 75 | 5.40E-47 | 6.45E-40 | 2.92E-08 | 3.48E-01 | 6.52E-01 |
| ENSG00000169174 | PCSK9 | 1 | 55039447 | 55064852 | 9.98 | 1.80E-23 | 8.41 | 4.06E-17 | 0.66 | < 2E-16 | 33 | 2.05E-25 | 2.17E-18 | 2.56E-10 | 1.71E-03 | 9.98E-01 |
| ENSG00000160712 | IL6R | 1 | 154405193 | 154469450 | 7.72 | 1.17E-14 | -7.84 | 4.36E-15 | -0.89 | < 2E-16 | 55 | 0.00E+00 | 0.00E+00 | 1.55E-09 | 1.00E+00 | 2.20E-04 |
| ENSG00000158710 | TAGLN2 | 1 | 159918107 | 159925507 | 3.02 | 2.54E-03 | 3.71 | 2.09E-04 | 0.84 | < 2E-16 | 26 | 3.11E-46 | 2.76E-43 | 4.08E-06 | 2.64E-03 | 9.97E-01 |
| ENSG00000083937 | CHMP2B | 3 | 87227271 | 87255556 | 3.47 | 5.15E-04 | 3.27 | 1.08E-03 | 0.62 | < 2E-16 | 290 | 7.83E-267 | 1.33E-258 | 5.88E-09 | 1.00E+00 | 3.28E-05 |
| ENSG00000165609 | NUDT5 | 10 | 12165330 | 12195891 | 3.58 | 3.37E-04 | 4.24 | 2.19E-05 | 0.52 | < 2E-16 | 40 | 1.48E-05 | 1.11E-03 | 1.42E-03 | 1.05E-01 | 8.93E-01 |
| ENSG00000140105 | WARS | 14 | 100333790 | 100376805 | -3.56 | 3.67E-04 | 3.67 | 2.42E-04 | -0.98 | < 2E-16 | 342 | 0.00E+00 | 9.57E-265 | 1.50E-87 | 1.00E+00 | 1.01E-05 |
| ENSG00000243678 | NME2 | 17 | 51165435 | 51171744 | -4.39 | 1.12E-05 | -3.3 | 9.63E-04 | 0.84 | < 2E-16 | 442 | 7.19E-303 | 1.67E-251 | 4.31E-52 | 1.00E+00 | 3.33E-32 |
| ENSG00000182534 | MXRA7 | 17 | 76672551 | 76711004 | -3.91 | 9.28E-05 | -3.27 | 1.06E-03 | 0.20 | < 2E-16 | 267 | 3.10E-296 | 1.09E-179 | 2.84E-117 | 1.00E+00 | 2.34E-92 |
| ENSG00000240972 | MIF | 22 | 23894383 | 23895227 | -4.95 | 7.33E-07 | 4.75 | 2.07E-06 | -0.97 | < 2E-16 | 202 | 1.67E-157 | 4.95E-54 | 2.72E-104 | 8.08E-01 | 1.92E-01 |

Chr: chromosome number; PP: posterior probability.

Among them, three genes (*IL6R*, *MIF*, and *WARS*) demonstrated opposite directions of association in TWAS and PWAS. Significant correlations were found between predicted transcript and protein levels for all ten genes, with negative coefficients noted for *IL6R*, *MIF*, and *WARS* (Table 1). These findings suggest shared genetic risk between transcripts and proteins for these genes. The absolute correlation coefficients ranged from 0.2 for *MXRA7* to 0.98 for *WARS*, indicating varying degrees of shared genetic components across genes.

Furthermore, we compared Z-scores from risk factor association analyses for these 10 genes (Figure 2). Predicted transcripts and proteins for the same genes showed consistent association patterns with CAD risk factors. For example, both predicted gene and protein expression levels of *PCSK9* were significantly associated with CAD risk factors such as total cholesterol, LDL, and CRP, and they were moderately correlated with a coefficient of 0.66. These results suggest that while distinct genetic factors may regulate gene and protein expression, their effects on CAD likely operate through shared biological mechanisms.

**Figure 2.**
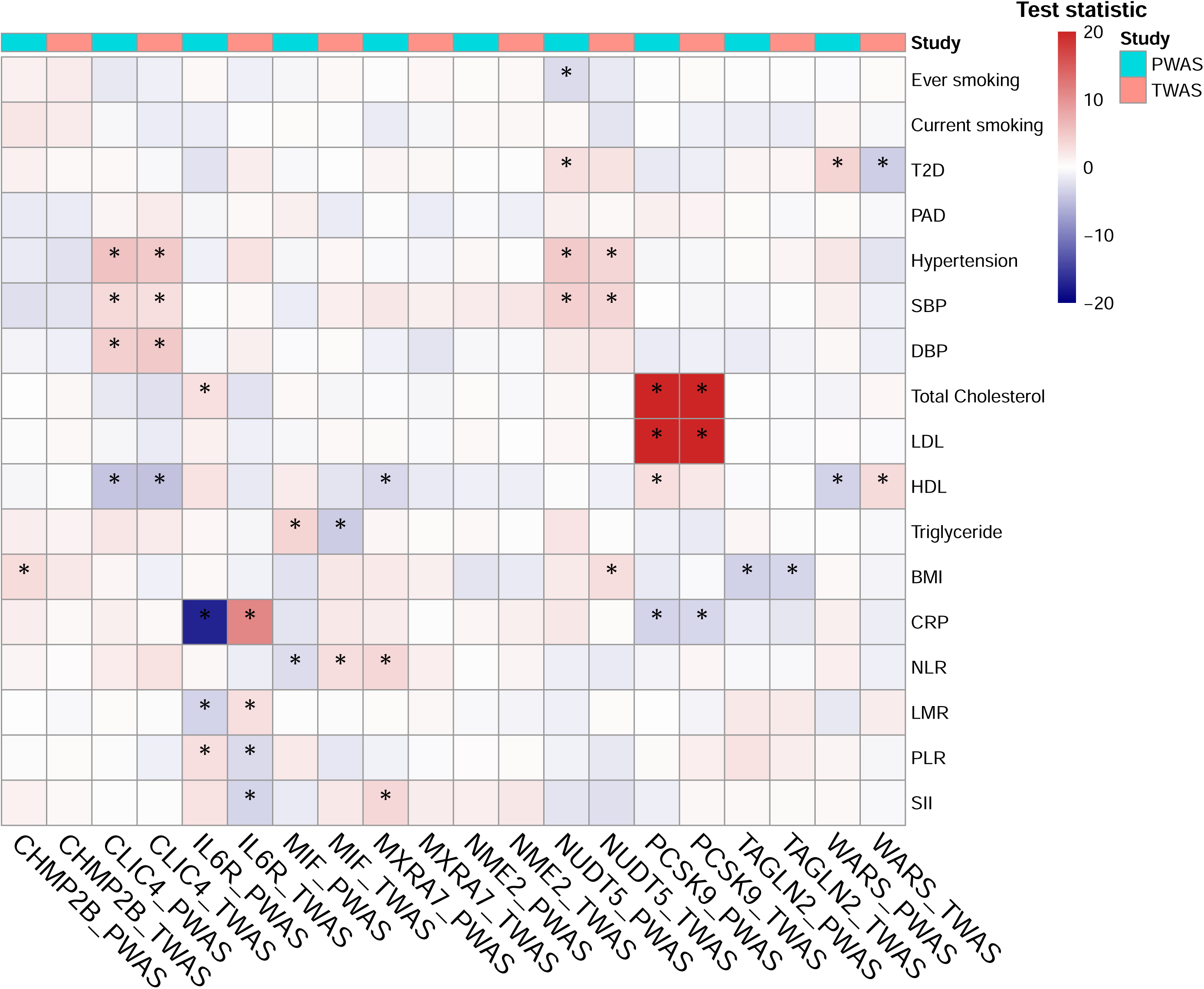
Associations between predicted transcripts and proteins of ten shared genes and CAD risk factors. Association results between the predicted transcripts and proteins of ten shared genes between TWAS and PWAS (columns) and CAD risk factors (rows). Each cell represents the test statistic (Z-score) of the association. P-value less than 0.05 was considered as significant (asterick). We noticed consistent patterns of associations for predicted transcripts and proteins of the same genes.

To explore the genetic basis of these correlations, we compared SNPs included in transcript and protein prediction models. Eight genes, excluding *CLIC4* and *MXRA7*, shared at least one common SNP (Supplemental Table 4). Of note, shared SNPs for *IL6R*, *MIF*, and *WARS* displayed opposite effect directions, consistent with the observed negative correlations.

We further conducted colocalization analyses to assess shared eQTLs and pQTLs for these genes. Four genes had PP4 greater than 0.6, while the remaining six showed PP3 values larger than 0.8 (Table 1). These results indicate that shared eQTLs and pQTLs for genes identified in both PWAS and TWAS may reflect common or correlated genetic risk factors.

## Discussion

We have developed a framework to analyze multi-omic risk profiles of diseases using GWAS summary statistics, transcript and protein prediction models, and individual-level genotype data from the UKB. Applying this framework to CAD, we identified 10 genes (*CHMP2B, CLIC4, IL6R, MIF, MXRA7, NME2, NUDT5, PCSK9, TAGLN2*, and *WARS*) for which both predicted transcript and protein levels were significantly associated with CAD. Significant correlations between predicted transcript and protein levels were observed for these genes. Analyses of model SNPs and colocalized shared QTLs revealed that, while the number of shared genetic signals varied among genes, the genetic components of gene expression and protein expression for the same gene likely contribute to CAD through shared molecular pathways.

There are several advantages of the proposed framework: First, integrating gene and protein expression data expanded our understanding of the genetic etiology of diseases by identifying additional disease-relevant genes.

Our findings significantly broaden the pool of risk genes for CAD in European populations. A comparison with the GWAS catalog revealed 113 unique genes without nearby SNPs previously reported to be associated with CAD. Notably, we identified a significant association between increased predicted expression of Actin-Related Protein 5 (*ACTR5*) and a reduced risk of CAD. Although no external studies currently support these findings, a recent study reported that suppressing *ACTR5* activates the expression of *CDKN2A*^22^, a gene located in the 9p21 chromosomal region, which is well-known for its association with CAD^23^. These results suggest that the *ACTR5-CDKN2A* pathway may represent a promising therapeutic target for CAD. Overall, our findings provide a foundation for future research on CAD-related genes and introduce potential new directions for investigating the genetic etiology of the disease.

Second, this framework enhances the interpretability of genetic risk factors by focusing on genes and pathways rather than individual variants.

For instance, our analysis identified an increased predicted expression of the gene *EIF6* as significantly associated with a decreased risk. Several nearby SNPs, such as rs6060235, have been reported in the GWAS catalog as relevant to CAD. However, prior studies have not mapped these SNPs to *EIF6*. While the specific mechanisms by which *EIF6* influences CAD are not fully understood, evidence suggests that it may function via the modulation of gut microbiota and inflammation^24^. Our analysis connected these known risk variants to a functionally relevant gene, offering new insights into the biological roles of these SNPs and their potential contribution to CAD development.

Third, integrating TWAS and PWAS provided valuable insights into disease mechanisms via molecular pathways, potentially aiding the development of targeted therapies.

We compared the results of TWAS and PWAS to identify 10 shared genes where both predicted transcripts and proteins were significantly associated with CAD risk. Of these, six genes (*IL6R, MIF, NME2, NUDT5, PCSK9*, and *WARS*) are established CAD risk genes. Our findings further validated their roles in CAD and provided insights into their potential biological mechanisms.

Existing evidence indicates negative relationships between genetic variants of interleukin-6 receptor (*IL6R*) and CAD^25,26^. Consistent with this, our PWAS results showed negative associations for *IL6R* protein expression, while the predicted transcript level was positively associated with CAD. This highlights the complexity of *IL6R* in CAD development, suggesting that specific *IL6R* variants may be protective against CAD while simultaneously increasing levels of soluble IL6R and IL6, both of which are pro-inflammatory factors^27^. Furthermore, our results imply that genetically predicted IL6R expression may influence CAD risk primarily through inflammatory pathways, such as elevating CRP levels, rather than directly impacting protein expression.

For four genes (*CHMP2B*, *CLIC4*, *MXRA7*, and *TAGLN2*), no nearby SNPs were associated with CAD. Mutations in *CHMP2B* are known to increase the risk of frontotemporal dementia^28^. While direct evidence linking CAD and frontotemporal dementia is lacking, dyslipidemia–a condition strongly associated with frontotemporal dementia–is a known risk factor for CAD^29^. Moreover, neurodegenerative diseases such as Alzheimer’s disease and dementia have been shown to share genetic risk factors with CAD^30,31^. These findings support the hypothesis that *CHMP2B* may influence CAD through shared pathways between neurodegenerative diseases and CAD.

There are some limitations in our current work. First, our analysis relies on established models that predict gene and protein expression levels from genetic variants, rather than direct measurements. The accuracy of these predictions is crucial to the validity of our findings. Furthermore, the predicted expression levels should not be considered equivalent to direct measurements due to variations in the heritability of gene and protein expression and the exclusion of non-genetic risk factors in the prediction models. Therefore, the observed associations should be interpreted as reflecting the relationships between the genetic components of omic levels and disease risk, rather than capturing the complete biological mechanisms.

Second, gene and protein expression levels vary across tissue and cell types, which may result in tissue-or cell type-specific associations with CAD. Thus, our findings based on whole blood may not be directly applicable to other tissues and all cell types. To assess this issue, we performed TWAS analyses using prediction models developed for coronary artery tissue and eight cell types on the same GWAS summary statistics.

Our results showed that most of the identified associations were robust to different tissues and cell type contexts. However, for PWAS, the lack of protein expression models for other tissues prevented us from conducting similar sensitivity analyses. Consequently, we emphasize that our results are derived from plasma-based analyses, and future studies should investigate these associations in other tissue types.

Third, our analysis assumes that mRNA levels directly influence protein expression levels of the same gene when interpreting the interplay between TWAS and PWAS findings. To validate this assumption, bivariate conditional analyses^8^ could be performed to assess whether the association between protein levels and disease risk is predominantly mediated by gene expression levels. However, given the central dogma of molecular biology and supporting evidence from the literature^32^, this assumption is unlikely to significantly affect the robustness of our findings.

Lastly, our study is restricted to genetically inferred European subjects due to the limited sample sizes of GWAS for other populations and the lack of gene and protein expression prediction models for non-European groups. Considering the differences in disease prevalence, heritability, and genetic architecture across diverse ancestry groups, these findings may not be generalizable to non-European populations. Future research will aim to incorporate larger datasets from non-European populations to address this limitation.

## Conclusions

We have developed a framework utilizing summary-level data to construct multi-omic risk profiles for diseases at the transcriptome and proteome levels, enhancing the understanding of genetic etiology. Applying this framework to CAD, we identified ten genes significantly associated with CAD in both TWAS and PWAS, highlighting potential molecular pathways involved in disease development. These findings expand the genetic risk factor pool for CAD and provide a robust foundation for future studies on CAD etiology, demonstrating the framework’s practicality and potential.

## Data Availability

The data that support the findings of this study are available from the corresponding author upon reasonable request.

## Acknowledgement

The authors would like to thank the researchers and participants of the United Kingdom Biobank. All data was accessed as part of project 32285 from the United Kingdom Biobank. We thank the Yale Center for Research Computing for the use of the McCleary High Performance Computing cluster.

## Supplemental Material

Tables S1-S4

Figure S1

## Non-standard Abbreviations and Acronyms

TWAS: Transcriptome-wise association study
PWAS: Proteome-wide association study
CAD: Coronary artery disease
GWAS: Genome-wide association study
ARIC: Atherosclerosis Risk in Communities
eQTLs: Expression quantitative trait loci
pQTLs: Protein quantitative trait loci
SNP: Single nucleotide polymorphism
MR: Mendelian randomization
UKB: UK Biobank
SBP: Systolic blood pressure
DBP: Diastolic blood pressure
TC: Total cholesterol
LDL: Low-density lipoprotein
HDL: High-density lipoprotein
TG: Triglycerides
BMI: Body mass index
CRP: C-reactive protein
NLR: Neutrophil-to-lymphocyte ratio
LMR: Lymphocyte-to-monocyte ratio
PLR: Platelet-to-lymphocyte ratio
SII: Systemic immune-inflammation index
PC: Principal component
FDR: False discovery rate
PP: Posterior probability

## Ethics approval and consent to participate

This research was conducted using the UK Biobank Resource (application number 32285). The UK Biobank study was conducted under generic approval from the National Health Services’ National Research Ethics Service. The present analyses were conducted in accordance with the Declaration of Helsinki and approved by the Human Investigations Committee at Yale University (2000026836 for UK Biobank). The patients/participants provided their written informed consent to participate in this study.

## Consent of publication

All authors agree to publish.

