## Supplementary material for "A framework for studying multi-omic risk factors and their interplay: application to coronary artery disease": ST4 and SF1

Supplemental Table 4. Shared SNPs in prediction models for eight genes.

| chr | pos | gene_name | rsid | ref_allele | eff_allele | weight_TWAS | weight_PWAS |
|---|---|---|---|---|---|---|---|
| 1 | 55052794 | PCSK9 | rs2495477 | A | G | -0.04 | -8.92 |
| 1 | 154449591 | IL6R | rs4845625 | T | C | -0.01 | 53.14 |
| 1 | 154453621 | IL6R | rs6689393 | A | G | 0.00 | 53.24 |
| 1 | 154453788 | IL6R | rs4129267 | C | T | 0.00 | 61.74 |
| 1 | 159922298 | TAGLN2 | rs2789422 | G | A | -0.01 | -15.09 |
| 1 | 159925746 | TAGLN2 | rs7513326 | G | A | 0.01 | 12.36 |
| 3 | 87214347 | CHMP2B | rs17024046 | C | T | 0.02 | 25.67 |
| 3 | 87255411 | CHMP2B | rs1060241 | G | A | 0.02 | 29.49 |
| 10 | 12207225 | NUDT5 | rs12773913 | G | T | -0.01 | -5.17 |
| 10 | 12207798 | NUDT5 | rs4747968 | C | T | -0.01 | -5.14 |
| 14 | 100337978 | WARS | rs7155068 | T | C | -0.03 | 32.34 |
| 14 | 100338949 | WARS | rs11624738 | C | A | -0.03 | 32.33 |
| 14 | 100370016 | WARS | rs941931 | C | T | -0.03 | 32.29 |
| 14 | 100370646 | WARS | rs4905956 | A | G | -0.03 | 32.29 |
| 17 | 51152947 | NME2 | rs16949649 | T | C | -0.01 | -29.35 |
| 17 | 51153539 | NME2 | rs2302254 | C | T | 0.00 | -28.19 |
| 17 | 51161675 | NME2 | rs2159359 | C | A | -0.03 | -28.17 |
| 22 | 23898153 | MIF | rs5760093 | G | A | 0.02 | -10.72 |

Chr: chromosome number; pos: positions.

Supplemental Figure 1. Quantile-quantile plots for TWAS and PWAS.

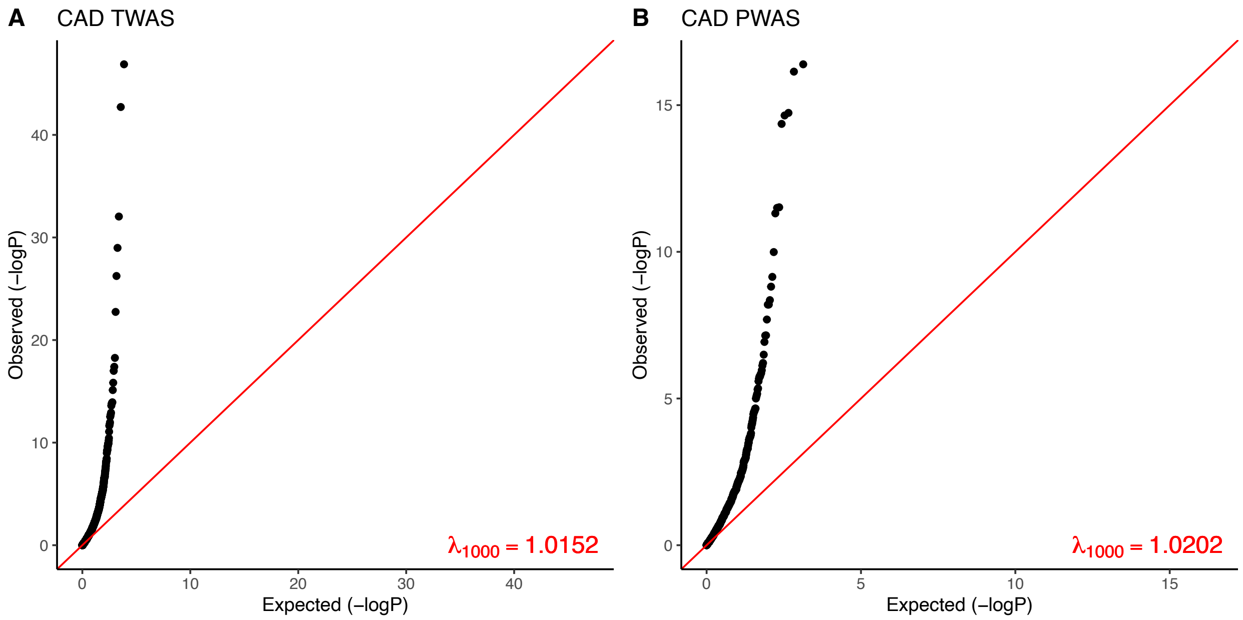


Quantile-quantile (QQ) plots for TWAS and PWAS results with $\lambda_{1000}$. Both TWAS and PWAS showed stable results with $\lambda_{1000}$ close to 1.05.
